# Personalized coronary and myocardial blood flow models incorporating CT perfusion imaging and synthetic vascular trees

**DOI:** 10.1101/2023.08.17.23294242

**Authors:** Karthik Menon, Muhammed Owais Khan, Zachary A. Sexton, Jakob Richter, Koen Nieman, Alison L. Marsden

## Abstract

Computational simulations of coronary artery blood flow, using anatomical models based on clinical imaging, are an emerging non-invasive tool for personalized treatment planning. However, current simulations contend with two related challenges – incomplete anatomies in image-based models due to the exclusion of arteries smaller than the imaging resolution, and the lack of personalized flow distributions informed by patient-specific imaging. We introduce a data-enabled, personalized and multi-scale flow simulation framework spanning large coronary arteries to myocardial microvasculature. It includes image-based coronary models combined with synthetic vasculature for arteries below the imaging resolution, myocardial blood flow simulated using Darcy models, and systemic circulation represented as lumped-parameter networks. Personalized flow distributions and model parameters are informed by clinical CT myocardial perfusion imaging and cardiac function using surrogate-based optimization. We reveal substantial differences in flow distributions and clinical diagnosis metrics between the proposed personalized framework and empirical methods based on anatomy; these errors cannot be predicted a priori. This suggests virtual treatment planning tools would benefit from increased personalization informed by emerging imaging methods.

## I. Introduction

Coronary artery disease (CAD) is the leading cause of death worldwide for both men and women [75, 54]. It causes narrowing or occlusion of coronary arteries, which can lead to myocardial infarction and loss of contractile heart function. Although clinical interventions such as stenting or coronary artery bypass graft (CABG) surgery can restore coronary flow, current long-term treatment outcomes remain unsatisfactory. While CABG improves mortality and morbidity in patients with severe narrowing or occlusion [87, 85, 80], 24% of patients report chest pain within a year [3], indicating sub-optimal perfusion even after treatment. Moreover, graft failure is a significant concern, with 50% of vein grafts exhibiting significant stenosis within 10 years after surgery [42]. There is strong evidence through large-scale randomized trials that clinical interventions guided by hemodynamic functional metrics, such as fractional flow reserve (FFR) [51, 73] and myocardial perfusion imaging [12, 7, 11, 46], lead to improved outcomes, lower rates of unnecessary revascularization, and lower costs [73, 52, 76, 1, 18, 33]. This is likely because anatomical imaging to guide the assessment of CAD severity and intervention correlates poorly with the functional severity of stenoses and myocardial ischemic risk [37, 38].

Computational fluid dynamics (CFD) simulations of coronary blood flow using anatomical image-based models are an emerging non-invasive method for risk assessment and treatment planning [36]. Clinical trials involving the FDA-approved FFR*_CT_* methodology, which uses patient-specific CFD to non-invasively compute FFR [72], have demonstrated increased diagnostic accuracy compared to anatomical imaging, a reduction in invasive catheterization procedures and increased cost-effectiveness in CAD risk stratification [30, 40, 25]. Patient-specific simulations of CABG patients have also produced biomechanical stimuli data, such as wall shear stress, that correlate with CAD progression and have shed light on mechanisms of vein graft failure [41, 57, 56, 55, 27, 58, 4]. Thrombotic risk assessment in Kawasaki disease has also demonstrated improved predictions using patient-specific models compared to standard anatomic metrics [21, 39].

However, the accuracy of patient-specific coronary flow simulations depends on the quality of imaging upon which they are based. A source of error often ignored in current computational models is the exclusion of arteries in the coronary anatomy that are under-resolved by imaging. CFD models are commonly constructed using coronary computed tomography angiography (CCTA) imaging which cannot accurately resolve vessels < 2 mm diameter [69]. This includes most vessels after the second generation of branching in the coronary artery tree [24]. Unsurprisingly, incomplete coronary artery branching in CFD models causes erroneous flow distributions amongst the arteries [82, 79]. Coronary flow distribution is also affected by the boundary conditions specified at each arterial outlet. A widely used method for prescribing these boundary conditions is Murray’s law [43, 88], which provides an empirical relationship between the flow through each artery and its diameter. However, this method depends purely on artery diameters, and does not account for occluded and collateral arteries, microvascular dysfunction, metabolic regulation or anatomical uncertainty. By ignoring inter-patient variability, this method is not personalized to each patient. As expected, these uncertainties in anatomy and boundary conditions significantly affect model predictions [78, 2, 62, 60, 34, 67, 32].

In this work, we propose a framework for improved accuracy of personalized coronary hemodynamics computational models that leverages: (1) recent advances in clinical imaging of myocardial perfusion to obtain personalized flow distributions in the coronary artery tree; (2) computational methods to model the anatomy and flow in microvasculature beyond the limits of clinical imaging resolution; and (3) data-driven tools to estimate personalized model parameters based on patient-specific clinical measurements. This framework is integrated into a simulation pipeline that reflects the multi-scale nature of the coronary circulation – including three dimensional flow in the large epicardial coronary arteries that can be reconstructed from CCTA, synthetic vascular trees for the small arteries that are under-resolved in clinical imaging, microvascular/capillary flow through the myocardial tissue, and finally closed-loop lumped parameter models for the four heart chambers and the distal/systemic circulation. In the proposed framework, we estimate patient-specific coronary flow distributions by incorporating non-invasive clinical imaging of myocardial perfusion. Dynamic CT myocardial perfusion imaging (MPI_CT_) is a CT-based non-invasive technique to quantify myocardial blood flow distribution [11, 46]. Imaging the coronary anatomy and myocardial perfusion can be performed using a single CT imaging modality, and this has been used to identify functionally significant CAD [1, 18, 33, 47]. Since the myocardium is perfused by coronary flow, the myocardial flow distribution can be used to estimate coronary flow distributions. This allows us to make personalized estimates of boundary conditions derived from clinical imaging which are not dependent on empirical rules or local vessel diameters [86].

Our framework also takes advantage of recently developed methods for computationally generating physiologically realistic vascular trees [63]. This allows us to create more physiological coronary artery anatomical models, combining CT-based anatomic models of the larger epicardial vessels, with synthetic vasculature for the distal vessels. While similar hybrid approaches have been introduced in previous studies [24, 49, 29], they have not yet incorporated high-fidelity 3D flow simulations, closed-loop circulation models, or personalized boundary condition estimation. Here, we demonstrate a novel technique to tune such vascular trees, as well as the parameters of closed-loop lumped parameter circulatory networks, based on patient-specific clinical measurements.

Finally, our hybrid image-based and synthetic vascular models of blood flow in the coronary arteries are coupled to downstream models of flow in the myocardial tissue. We model myocardial perfusion at microvascular/capillary scales as flow through a porous material, following previous studies [5, 9, 22, 13, 49, 29]. Such models are dictated by a several parameters, including the properties of the tissue and the coupling between the myocardial blood flow and the upstream flow in the coronary artery tree. While previous studies have discussed the parameterization and estimation of tissue properties [22, 14], they have not personalized the distribution of myocardial perfusion – which is affected by the upstream coronary arteries. The focus of the current work is therefore to recapitulate clinically measured myocardial perfusion using a multi-scale model of coronary flow.

The overarching aim of this work is to build accurate and personalized computational models that recapitulate clinically measured coronary hemodynamics and myocardial perfusion. Such models provide a crucial baseline for performing virtual treatment planning in future work. We therefore describe the framework in the context of patient-specific data acquired prior to CABG surgery. The objectives and novelty of this paper are hence as follows: (1) we demonstrate a personalized multi-scale computational simulation pipeline for coronary flow from the large blood vessels to the capillaries in the myocardium, which is informed by MPI_CT_; (2) we demonstrate that parameter estimation can effectively and efficiently recapitulate pre-treatment clinical measurements; (3) we compare hemodynamics of these personalized models with those constructed using conventional Murray’s law-based boundary conditions and without synthetic vascular trees to highlight the importance of model personalization.

## II. Methods

### i. Patient population

We performed simulations on a subset of 4 patients (figure 1) from a cohort who were scheduled to undergo coronary artery bypass graft (CABG) surgery, and also underwent coronary CT angiography (CCTA) and dynamic CT myocardial perfusion (MPI_CT_) imaging at Stanford University School of Medicine, Stanford CA, USA as part of an ongoing study (NCT03894423). This study was approved by the Institutional Review Board at Stanford University School of Medicine. Written informed consent was received prior to patient participation. The exclusion criteria for the cardiac CT exam were: (1) Age<40 years; (2) left ventricular ejection fraction <30%; (3) repeat CABG; (4) contra-indications to iodine contrast medium, including eGFR<45 ml/min, known contrast allergy; (5) contra-indications to vasodilators, including bronchial asthma, advanced atrioventricular block, sinus node disease, clinically significant carotid artery narrowing, severe aortic stenosis or left ventricle outflow tract narrowing, blood pressure <90mmHg, use of dipyridamole or aminophylline; (6) conditions technically challenging the examination, including severe valvular disease, atrial fibrillation and body weight >100kg; (7) other conditions of that would render the examination unsafe, including pregnancy, unstable ischemia, severe arrhythmia, heart failure. From the patient cohort that underwent a cardiac CT exam, the inclusion criteria for this computational modeling study were the availability of CCTA and MPI_CT_ images of sufficient quality to re-construct the 3D coronary anatomy and myocardial blood flow (MBF), as well as clinical measurements of heart rate, blood pressure, cardiac output and ejection fraction measured at the time of the CCTA.

**Figure 1:**
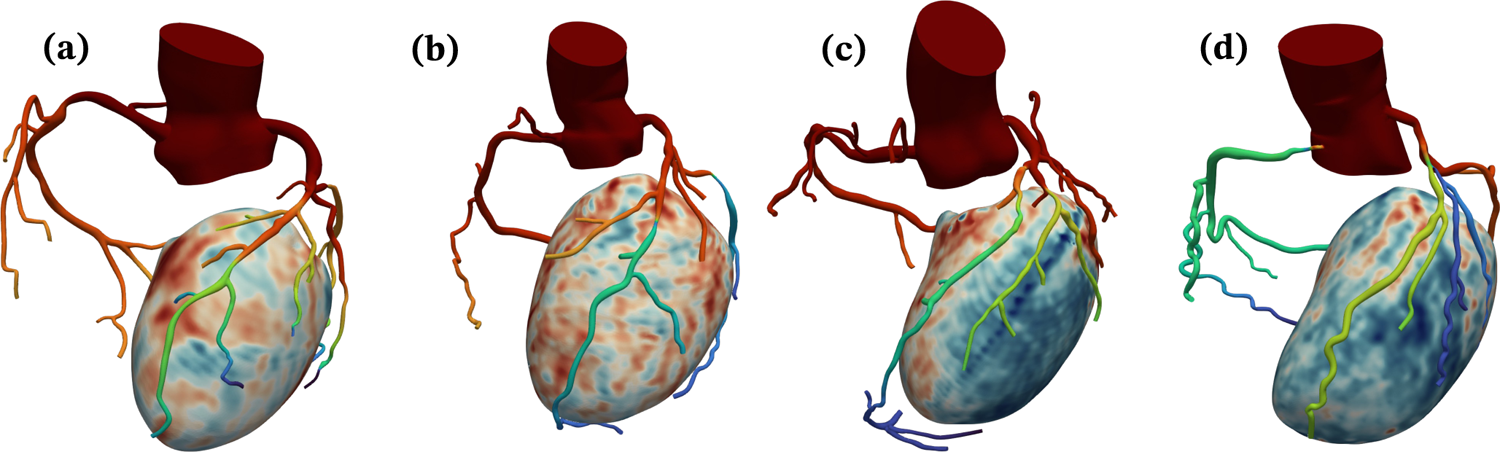
Coronary and LV anatomical models for the four patients in this study. The colors on the coronary arteries qualitatively represent simulated average pressure over a cardiac cycle and the colors on the LV represent MBF from MPI_CT._ Figures (a), (b), (c) and (d) show patients 1, 2, 3 and 4, respectively.

### ii. Clinical imaging protocols and processing

MPI_CT_ followed by CCTA were performed on a third-generation dual-source CT scanner (SOMATOM Force, Siemens Healthineers). Hyperemia was induced by slow bolus injection of 0.4 mg regadenoson. A 40-ml bolus of iopamidol (Isovue, 370 mg/ml, Bracco, Italy) was injected at 5.5 ml/s followed by 40 ml saline. The dynamic perfusion protocol requires that the table position is repeatedly alternated (shuttle mode) to completely cover the myocardium and acquire up to 15 data samples during a 30s breath hold. Acquisition parameters were 80-kV tube voltage, automated exposure control (300 mAs/rotation at 80 kV as reference), 96×0.6-mm detector collimation, 105-mm z-axis coverage, 250-ms gantry rotation time, and 66-ms temporal resolution. Reconstructed images with a 3.0-mm slice thickness, and 2.0-mm reconstruction interval were processed on a dedicated workstation (Syngo.CT Myocardial Perfusion, Siemens Healthineers). Serial samples were aligned using a motion correction algorithm. The arterial input function was sampled in the descending aorta. Time-attenuation curves were created for each voxel within the segmented left ventricle (LV) myocardium. Using a dedicated parametric deconvolution algorithm based on a 2-compartment model of intra- and extravascular space, a fit curve was created. MBF was calculated as the ratio between the maximum slope of the fit curve and the peak arterial input function, and the results were exported as three-dimensional MBF maps (figure 2a). Comprehensive descriptions of the myocardial perfusion protocol and the calculation of MBF can be found in [35].

**Figure 2:**
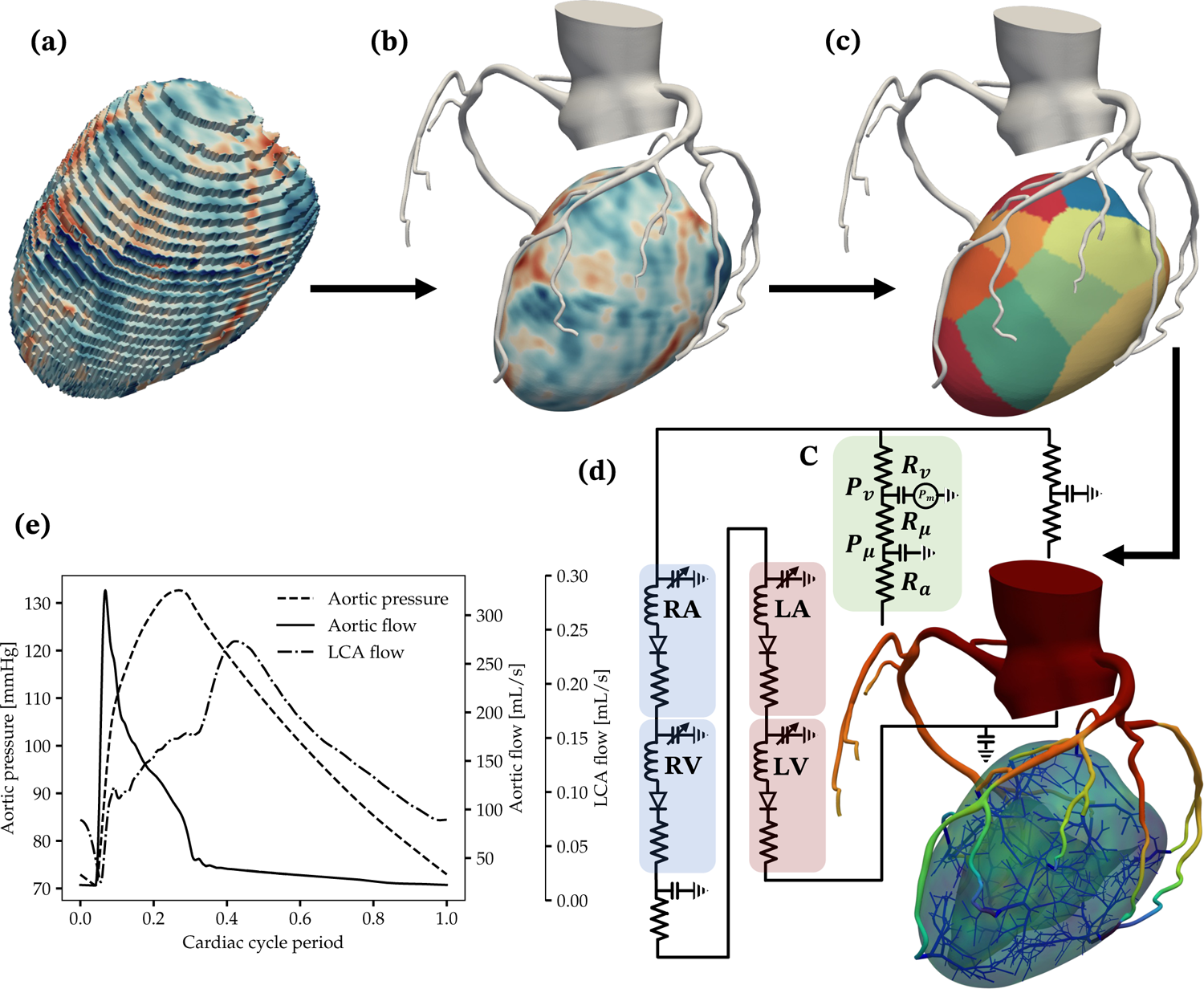
Overview of the computational modeling workflow. (a) Raw volume mesh of the LV, with color contours showing MBF from MPI_CT_. (b) Segmented image-based coronary tree and the LV models after co-registration and processing. (c) LV perfusion volumes corresponding to each coronary artery. (d) The flow simulation setup, consisting of 3D models of the coronary arteries and LV, synthetic vascular trees perfusing the LV, and closed-loop boundary conditions. RA, RV, LA, LV and C denote the right atrium, right ventricle, left atrium, left ventricle, and coronary outlet LPN models respectively. (e) Simulated aortic pressure, aortic flow and left coronary flow over a cardiac cycle.

A standard CCTA was performed 10 minutes after MPI_CT_ by prospective ECG-triggered axial scan mode. The tube voltage ranged between 100 and 120 kV. Most patients received 50mg aminophylline after the perfusion scan, as well as intravenous metoprolol if the heart rate exceeded 70 bpm. All patients received sublingual nitroglycerin just before the contrast-enhanced CT exam.

Since the CCTA and MPI_CT_ were performed at mid-diastole and systole respectively, they were co-registered using the affine registration procedure implemented in *3D Slicer* (www.slicer.org). The LV myocardial volume was then segmented from the co-registered MPI_CT_ image using thresholding in *ParaView (*www.paraview.org). It was smoothed and meshed with tetrahedral elements using *Meshmixer (*www.meshmixer.com) and *TetGen* [70], respectively. Finally, the MBF values at each voxel of the MPI_CT_ scan were interpolated on to the nodes of the LV mesh. Note that all the software used for the processing of these images are freely available, and all except *Meshmixer* are open-source. Figure 2(b) shows the 3D volumes of the co-registered LV and coronary vasculature for patient 1.

Following the construction of co-registered 3D models of the LV myocardium and coronary arteries, the LV volume was divided into non-overlapping sub-volumes corresponding to the perfusion territory of each coronary artery (figure 2c). This was done using Voronoi tessellation, where each node in the tetrahedral mesh for the LV volume was assigned to its closest coronary artery outlet. Therefore, each coronary artery outlet was associated with a distinct region of the LV myocardium. Subsequently, the flow-rate associated with each coronary outlet was estimated by integrating the MBF obtained from MPI_CT_ within each corresponding LV perfusion territory. We note that the LV perfusion territories corresponding to each coronary artery can also be computed based on the distance of each point in the LV to the closest coronary artery (instead of coronary artery outlet) [62, 86] as well as weighted measures of distance [13, 49]. This approach was chosen for simplicity, however, the framework presented in this work is equally applicable to any method for computing perfusion regions in the LV.

### iii. Image-based modeling and coronary flow simulations

We used the open-source *SimVascular* software [77] for segmentation of coronary arteries from CT angiography images, construction of 3D anatomical models, and coronary flow simulations. Tetrahedral finite element meshes were created using the open-source *TetGen* package [70], which is included with *SimVascular*. Three dimensional flow simulations were performed using the *svSolver* flow solver within *SimVascular*, which uses a stabilized finite element method with linear tetrahedral elements for spatial discretization and generalized-*α* time-stepping with second-order time accuracy [23, 83].

The governing equations for coronary blood flow were the three-dimesional incompressible Navier-Stokes equations,

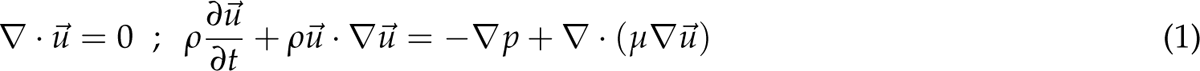

where ⃗*u* and *p* are the blood flow velocity and pressure, respectively. Blood was assumed to be a Newtonian fluid with viscosity *µ* = 0.04 dynes/cm^2^ and density *ρ* = 1.06 g/cm^3^. The Newtonian fluid assumption is valid because non-Newtonian effects begin to be apparent only for blood vessels with diameters below 300 *µ*m [65], which was approximately the limit of the smallest vessels modeled in this study.

Coronary artery walls were treated as deformable, and fluid-structure interaction arising from the coupling between blood flow and the artery walls was handled using the coupled momentum method [16]. Material properties for the artery walls were selected based on literature data following previous work [20, 39]. The elastic modulus for the aorta and coronary arteries were set to 0.25 MPa and 1.15 MPa, respectively [19, 8, 59]. Wall thicknesses were based on published radius-thickness ratios for the aorta [8] and morphometric data for the coronary arteries [53].

### iv. Synthetic vascular trees

We augmented our image-based coronary artery anatomical models with synthetic vascular trees to model coronary vasculature with diameters below the imaging resolution of the CCTA. Constrained Constructive Optimization (CCO) [63] was used to generate synthetic vascular trees that are constrained by vascular scaling laws and hemodynamic factors while minimizing vascular volume. CCO has recently been adapted to generate multiple non-intersecting trees within non-convex ventricle-like tissue volumes [24], although this process is computationally expensive. In this work we used an improved open-source implementation of Sexton *et al.* [68] to more efficiently generate synthetic vascular trees at the outlet of each coronary artery that perfused the LV. While the CCO implementation used here allows the generation of several competing vascular trees within the same tissue volume, we chose to generate one independent vascular tree within the LV volume associated with each coronary artery. In our experience, this approach resulted in the LV being more uniformly vascularized and with significantly lower computational cost by avoiding the generation of unphysiological competing tree morphologies from approximately 15 coronary artery outlets.

For each case simulated in this work, we generated synthetic vasculature with a total of 500 outlet branches perfusing the LV (compared to approx. 16 branches perfusing the LV in the purely image-based anatomical models). The total number of synthetic outlet branches was divided amongst the vascular trees appended to each coronary artery outlet based on the size of their corresponding LV perfusion volumes. The smallest vessels produced were approximately 300 *µ*m in diameter. Flow in the synthetic vasculature was modeled using a lumped-parameter/0D representation of the vessels, which included resistance, capacitance and inductance to model viscus losses, wall dilation, and fluid inertia respectively. The lumped-parameter resistance, capacitance and inductance of each vessel were given by linearization and Poiseuille flow simplifications of the Navier Stokes equations [50]. The lumped-parameter flow in these synthetic vascular trees was simulated using the open-source *svZeroDPlus* solver, which is part of the *SimVascular* software suite, and was coupled to the hemodynamics in the 3D image-based anatomical model using a modular implicit coupling method [15].

### v. Coronary flow boundary conditions

We used a closed-loop lumped parameter network (LPN) model to enforce boundary conditions for the coronary flow simulations at the aortic inlet and the aortic and coronary outlets of the 3D model [61, 74, 39] (figure 2d). The LPN included specific features to model distal systemic resistances using Windkessel models [81] and the effect of the four heart chambers. It also captured the distal resistance and intra-myocardial pressure experienced by coronary arteries, which produces out-of-phase coronary flow with respect to the cardiac cycle [28]. Figure 2(e) shows representative pressure and flow waveforms for one case. As seen in figure 2(d), the LPN boundary conditions at the coronary artery outlets consisted of three resistance elements – *R_a_*, *R_µ_* and *R_v_*, to model the resistance of small arteries, microvasculature, and the venous system respectively. These resistance and capacitance elements distal to each coronary artery in the model, as well as other parameters of the closed-loop LPN, such as the heart chamber elastances and capacitances, and systemic resistances, were estimated to match clinically measured data (see section vii). As with the lumped parameter representation of the synthetic vascular trees discussed in section iv, the closed-loop LPN boundary conditions were simulated using *svZeroDPlus* and coupled with the 3D flow solver using a modular implicit coupling method [15].

We also simulated coronary flow at hyperemia to recapitulate common clinical measurements that assess coronary hemodynamics under stress (including FFR and MPI_CT_). We simulated hyperemia by scaling the resistance boundary conditions distal to coronary arteries by a factor of 0.24 compared to baseline resting values [84], as commonly done in FFR_CT_ [72].

### vi. Myocardial blood flow simulations

Myocardial perfusion in the arterioles and capillaries within the LV myocardium was modeled as flow through a porous material due to the prohibitive computational cost of resolving discrete microvascular blood vessels [5, 9, 22, 13, 49, 29]. Flow through porous materials is described by Darcy’s Law, and in this work we used a single-compartment Darcy model governed by the equations,

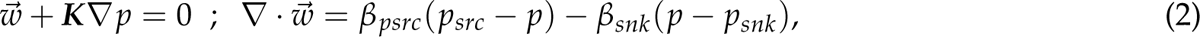

where *w*∼ and *p* are the Darcy flow velocity and pressure, respectively. The permeability of the LV myocardial tissue is given by ***K***. We assumed constant and isotropic permeability for simplicity, as in prior Darcy models for myocardial perfusion [5, 13, 14, 49], i.e. ***K***(⃗*x*) = *k**I***, where ***I*** is the identity tensor and *k* is the scalar, spatially-constant permeability. We fixed *k* = 2 × 10*^−^*^5^ cm^2^ Pa*^−^*^1^ s*^−^*^1^, which is informed by previous work [5, 49]. Myocardial perfusion was driven by the coronary flow in the upstream vasculature via pressure source terms, which are denoted by *p_src_* in equation 2. These pressure sources, corresponding to the outlet of each coronary artery that is perfusing the LV, were specified as the microvascular pressure in the coronary outlet boundary condition model (see section v and *P_µ_* in figure 2d). Therefore, each perfusion territory associated with a coronary artery outlet, discussed in section ii, was assigned a corresponding pressure source. The pressure of the venous return system is given by *p_snk_* in equation 2, and represents a sink for myocardial perfusion. We assumed *p_snk_* = 0 mmHg in this work [49]. Lastly, the terms *β_src_* and *β_snk_* represent the conductance of flow entering and leaving the tissue, respectively, and were parameterized as follows,

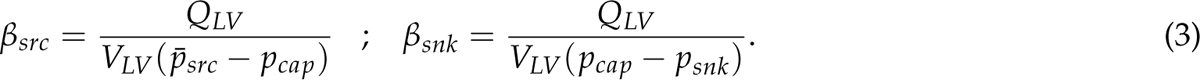

In equation 3, *Q_LV_* and *V_LV_* are the total flow perfusing the LV and the volume of the LV respectively. The spatially-averaged pressure source (*p_src_*) is denoted as *p*^-^*_src_* and *p_cap_* is the target capillary pressure assumed to be *p_cap_* = 15 mmHg [5, 49]. Note that for a given tissue volume *V_i_ V_LV_*, such as one vessel-specific perfusion territory, the MBF within that volume is given by,

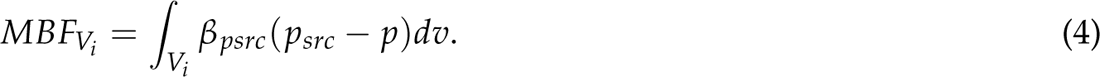

We simulated perfusion in patient-specific LV volumes segmented from MPI_CT_, as described in section ii. Unstructured tetrahedral meshes were generated using the open-source *TetGen* package [70], and pressure sources (*p_src_*) were imposed from flow simulations in the coronary arteries for each patient-specific case. A finite element solver for equation 2 employing linear tetrahedral elements was implemented in the open-source *svFSI* software, which is part of the *SimVascular* software suite. We used zero-flux boundary conditions on the surface of the LV mesh.

### vii. Parameter estimation for personalized boundary conditions

We employed a multi-stage surrogate-based optimization procedure to estimate the parameters of the closed-loop LPN boundary condition model (discussed in section v) to recapitulate measured clinical data for each patient. This was performed for purely image-based models as well as those augmented with synthetic vascular trees. The clinical targets we used were the vessel-specific flows from MPI_CT_; systolic and diastolic aortic pressure from blood pressure cuff measurements; left ventricular ejection fraction, stroke volume, cardiac output and systolic/diastolic ventricular volumes from echocardiography; and physiological targets derived from literature including pulmonary pressure and coronary flow waveforms [31, 48, 74]. We note here that the vessel-specific flow and MBF targets from MPI_CT_ were calculated relative to the total LV flow because MPI_CT_ is known to underestimate the absolute flow [46, 86]. As a basis for comparison, we also tuned each model to match all the above clinical targets except vessel-specific flows. In these latter models, the distribution of flow amongst the coronary arteries was prescribed based on their lumen diameters using Murray’s law [43, 88], as is more conventionally done.

The surrogate model we used for the optimization was a lumped parameter representation of each patient-specific coronary artery anatomical model. This was generated using an automated method [50] that extracts the lumen centerlines of all vessels in the anatomy, computes vessel radii by traversing along the centerlines, and distills this information into a network of resistance-capacitance-inductance lumped parameter models for each vessel in the anatomy. As in the lumped-parameter representation of the synthetic vascular trees discussed in section iv, the resistance, capacitance and inductance of each vessel were given by linearization and Poiseuille flow simplifications of the Navier Stokes equations [50]. We henceforth refer to this as a 0D surrogate model.

### vii.1 Optimizing for cardiac function

In the first stage we optimized the parameters of the closed-loop system to match all the above clinical targets except the vessel-specific flows. This consisted of 36 parameters governing the hemodynamics and function of the four heart chambers, the pulmonary circulation, the intramyocardial pressure experienced by coronary arteries, and the total distal resistance and capacitance at all the aortic and coronary outlets. For details about the parameters optimized in this stage, see reference [39]. This optimization was performed using the Nelder-Mead method, a deterministic gradient-free optimization technique [45]. The convergence tolerance for the optimization was 0.1, maximum number of iterations was 200, the parameter update step size was 0.1 of the range for each parameter, and the number of restarts was 50.

The initial guess for the total vascular resistance for each patient was determined from the clinically measured average aortic pressure divided by cardiac output. The total coronary flow was given by the morphometric scaling with the LV volume, *Q_LV_* = 3.41 *V*^0.75^ mL/min for branches perfusing the LV and 0.2 × *Q_LV_* for other branches [6, 10, 49, 86]. For the models tuned to match the flow distribution measured by MPI_CT_, the total coronary resistance was distributed amongst the branches of the coronary tree based on the ratio of total MBF to vessel-specific MBF for each vessel. On the other hand, for the models tuned to match flow distributions resulting from Murray’s law, the total coronary resistance was distributed amongst all coronary branches based on their diameters scaled to an exponent of 2.6 [88]. The total capacitance was set as 0.28 10*^−^*^3^ cm^5^/dynes for aortic branches and 10*^−^*^6^cm^5^/dynes for coronary branches based on previous work [61]. The total capacitance was distributed amongst branches to be proportional to their outlet areas [6]. When tuning all the models, the permissible range for each parameter was assigned based on physiological values and the initial guess was based on the maximum posterior of the distribution of parameters computed using Markov Chain Monte Carlo techniques in our previous work using similar closed-loop LPN models for coronary hemodynamics [74].

### vii.2 Optimizing for blood flow distribution: Without synthetic vasculature

The second stage of optimization focused on vessel-specific outlet boundary conditions. In this stage, we kept the 36 parameters that were optimized in the first stage fixed, and tuned the outlet resistances (represented by *R_a_*, *R_µ_* and *R_v_* in figure 2(d)) of all the branches in the coronary tree to match clinically measured flow distributions from MPI_CT_.

For the models that were not augmented with synthetic vascular trees, this consisted of two parameters for each of the *N_outlets_^CT^* coronary outlets perfusing the LV – the total outlet resistance for each branch (*R^i^_a_* + *R^i^_µ_* + *R^i^_v_*, where *i = 1,…, N^CT^_outlets_* is the index of the outlet), and the ratio of proximal vessel to microvascular resistance at each outlet (*R^i^_a_*/*R^i^_µ_*). The former determines the flow distribution in the coronary tree while the latter dictates the distribution of microvascular pressure that drives myocardial perfusion. We first used Nelder-Mead optimization to tune the relative distribution of total resistance amongst all the coronary artery outlets so that the resulting distribution of flow in the coronary tree matched that measured from MPI_CT_. This was done using a *N^CT^*_outlets_-dimensional optimization procedure that estimated a scaling factor constrained to the range [0.5, 2.0] for the resistance at each outlet, such that the total outlet resistance of the model was kept equal to the value obtained from the first stage of optimization. The convergence tolerance for the error was set to 0.1, the maximum number of iterations was 200, the parameter update step size was 0.1 of the range, and the number of restarts was 50.

We then fixed the total resistance of the complete tree as well as the total resistance at each outlet, and estimated the ratio of proximal to microvascular resistance that matched the required distribution of microvascular pressure. Note that the required microvascular pressure source (*p_src_* in equation 2) in each vessel-specific perfusion territory can be computed from equation 4. This is given by *p^i^_src_* = *MBF_Vi_*/(*β_src_V_i_*) + *p_cap_*, where a guess for all the pressure sources is required to compute *p*^-^*_src_*in *β_src_*. In addition, the simulated microvascular pressure at each outlet *i* in the flow simulation is given by *P^i^_μ_* = *P^i^ − Q^i^ R^i^_a_*, where *P^i^* and *Q^i^* are the pressure and flow at outlet *i* (see figure 2d). We therefore ran a 3D simulation with deformable walls to estimate the outlet pressure *P^i^* and microvascular pressure *P^i^* at each coronary outlet. For this initial simulation, we assumed the ratio of proximal to microvascular resistance was *R^i^_a_*/*R^i^_μ_* = 0.38 at each outlet, following previous work [61, 39]. This computation of the pressure loss along each branch, combined with the required *p^i^* for each branch, informed a direct update of *R^i^_a_*/*R^i^_μ_* at each outlet to obtain the required microvascular pressure. This updated ratio of proximal to microvascular resistance, along with the optimized distribution of outlet resistances, was then used to run a 3D simulation with deformable walls at hyperemia. The microvascular pressure obtained at each outlet from this simulation, along with the required microvascular pressure source for each outlet from equation 4, was then used to inform a second update of the ratio between the proximal and microvascular resistance at each coronary outlet.

### vii.3 Optimizing for blood flow distribution: With synthetic vasculature

For the models augmented with synthetic vascular trees, a different approach was employed for tuning the boundary conditions at the outlets, since these models consisted of *N^syn^_outlets_* = 500 coronary outlets (compared to ~ 16 outlets without synthetic vasculature), rendering the approach above computationally intractable. Moreover, since the synthetic trees generated by CCO are not patient-specific, we noticed that the vascular resistance along some paths from the aorta to outlet vessels led to unphysiological outlet boundary conditions (such as negative resistance) in order to match clinically measured flow and pressure distributions. Therefore, we developed a novel iterative approach to personalizing synthetic vasculature that estimates not only the parameters at the coronary outlets, but also dilates or constricts the synthetic vessels to match patient-specific flow and pressure targets.

Each iteration of the procedure consisted of: (1) sub-iterations to match the required flow distributions in the 0D surrogate to within 5% error, which involved updating the distal resistance boundary condition at each outlet vessel and dilating the synthetic vascular trees if unphysiological negative resistances were required to match the flow targets; (2) a simulation of the updated surrogate model and comparison of the outlet pressure distribution with that required to match clinically imaged MBF; (3) the dilation or constriction of all the synthetic vascular trees to obtain the required pressure distribution if the average pressure error was greater than 5%. The above steps were iterated upon until both the flow and pressure errors (based on the 0D surrogate model) were below 5%. Each of these steps is described in detail below.

With the parameters derived from the first stage of optimization, we first ran a 0D simulation at hyperemia to compute the pressure at all outlets. Using the aortic pressure and outlet pressures, we estimated the linear vascular resistance along each path from the aortic inlet to a coronary outlet, given by *R^i^_path_* = (*P_aorta_ − P^i^*)/*Q^i^*, where *P^i^* and *Q^i^* are the pressure and flow at an outlet synthetic vessel indexed by *i*. We also computed the resistance of the paths from each outlet of the image-based epicardial vessels to all outlets of the synthetic tree that was appended to that epicardial vessel. This is given by *R^i^_path,syn_ = (P^i^_epi_ − P^i^)/Q^i^* where *P^i^^epi^* is the pressure at the outlet of the image-based epicardial vessel which is upstream of the synthetic vessel outlet *i*.

Given the resistance *R^i^_path_* along each aorta-to-outlet path and the total resistance of the model estimated from the first stage of optimization (*R_tot_*), we calculated the required resistance distal to each outlet as *R^i^_out,reqd_* = *R_tot_Q_LV_*/*Q^i^_reqd_ − R^i^_path_*, where *Q^i^_reqd_* is the target flow through outlet *i*, according to MPI_CT_. This informed an updated resistance distal to each outlet. For some outlets in the model, this estimated outlet resistance was negative. This was due to excessive pressure drop along the path from the aorta to outlet *i*, i.e. *R^i^_path_* was too large. Since each *R^i^_path_* consisted of the sum of resistance from the upstream image-based epicardial vessels and downstream synthetic vessels, we addressed this by dilating the synthetic vascular trees. This achieved the required resistance along each path while maintaining the anatomy of the upstream branches that were based on patient-specific imaging. For each *R^i^_out,reqd_* that was negative, we estimated the required resistance of the path from the outlet of the corresponding image-based vessel to the synthetic outlet *i*, which we denote as *R*^∼^ *^i^_path,syn_*, such that *R^i^_out,reqd_* > 0. Note that *R*^∼^ *^i^*, since we are aiming to decrease *R^i^_path_*. Each of these synthetic vessel outlets was then assigned a radius scaling factor to achieve this reduced resistance, given by *α = ^i^_path,syn_/*(*R^i^_path,syn_*)^1/4^ > 1. The synthetic trees were then dilated using the method described below. The above procedure was iteratively performed until physiological outlet resistances were attained for each synthetic vessel outlet and the average error in the outlet flow fractions, *ɛ_flow_* = ∑*_i_*(*|Q^i^_reqd_ − Q^i^_reqd_*)N^syn^_outlets_, was less than 5%.

We scaled the radii of synthetic vessels after each outlet vessel was assigned a radius scaling factor, *α^i^*, by traversing up each synthetic tree from the outlet vessels to the root of the tree. As we traversed up the tree, each parent vessel was assigned a scaling factor that was the maximum of the scaling factors of its two daughter vessels. Moreover, the maximum scaling factor for each vessel was constrained by the radius of its parent vessel. This ensured the preservation of the morphometric structure of the tree. In this way, each vessel in every vascular tree was assigned a scaling factor *α^i^*by propagating the outlet vessel scaling factors up the tree. We then updated the resistance, inductance and capacitance values of all the synthetic vessels based on the new radius.

Once the outlet boundary conditions were updated through the above procedure, we tuned the vascular resistance of the synthetic vasculature to obtain the required microvascular pressure distribution at the coronary outlets based on the MBF distribution from MPI_CT_. As discussed in section vii.2, the target microvascular pressure source at each coronary outlet (*p_src_* in equation 2) can be estimated from equation 4. This is given by = *MBF_Vi_* /(*β_src_V_i_*) + *p_cap_*. Therefore, the target pressure at every synthetic vessel outlet with index *i* can be calculated as *P^i^_reqd_ = p^i^_src_ + Q^i^_reqd_R^i^_a_*. Furthermore, the required vascular resistance along each path from the aortic inlet to each synthetic vessel outlet can be calculated from the required outlet pressure and required outlet. Therefore, we ran a second 0D simulation at hyperemia to estimate the pressure at all the synthetic vessel outlets. We computed the average relative pressure error over all the outlets as *∈_pressure_ = Σ_i_(|P^i^ − P^i^_reqd_~/P^i^_reqd_)N^syn^_outlets_*. If this error was greater than 5%, we proceeded as follows to update the resistance of synthetic vascular trees. The 0D simulation again allowed us to calculate the path-specific vascular resistance from the aortic inlet to each synthetic tree outlet (*R^i^_path_*). Similarly, we calculated the resistance from each image-based epicardial coronary outlet to the synthetic vessel outlets that were appended to that image-based vessel. Given the required resistance along each path computed above (*R^i^_path,reqd_*, and R^i^_path,syn_) the separate contributions to this resistance from the image-based vessels and synthetic vessels (*R^i^_path_*), we computed the required vascular resistance of the synthetic vessels along the path from the outlet of the corresponding upstream image-based vessel to the synthetic outlet *i*. This resistance, denoted by *R*^∼^ *^i^_path,syn_*, then informs a radius scaling factor) *α_i_ = (R^i^_path,syn_/R^i^_path,syn_)*^1/4^ for the each synthetic vessel outlet *i*. We constrict/dilate the synthetic vascular trees using the procedure described above, and then return to the start of the iteration to estimate new outlet resistances and the flow distribution resulting from this updated anatomy.

## III. Results

We applied the parameter estimation framework to personalize 4 patient-specific coronary hemodynamics models, as discussed in section i. The quality of the CCTA was higher for cases 1 to 3, compared to case 4, which was included to highlight the challenge posed by lower-quality clinical imaging. The imaging quality dictated the number of coronary artery branches that could be segmented for each anatomical model, resulting in an average of 16 coronary artery branches perfusing the LV in cases 1 to 3, and 9 branches for case 4. This section discusses the performance of the framework at recapitulating clinical measurements, as well as comparisons with boundary conditions based on Murray’s law.

The performance of the first stage of the model personalization framework, which focused on estimating the parameters of the closed-loop LPN heart circulation model, is shown in figure 3. The framework successfully recapitulated patient-specific clinical targets, comprised of systolic/diastolic blood pressure and echocardiographic measurements of ejection fraction and ventricle volumes, for all 4 patients. The average error between the simulated and clinically measured targets was 5.07% for patient 1, 6.84% for patient 2, 4.36% for patient 3 and 4.16% for patient 4.

**Figure 3:**
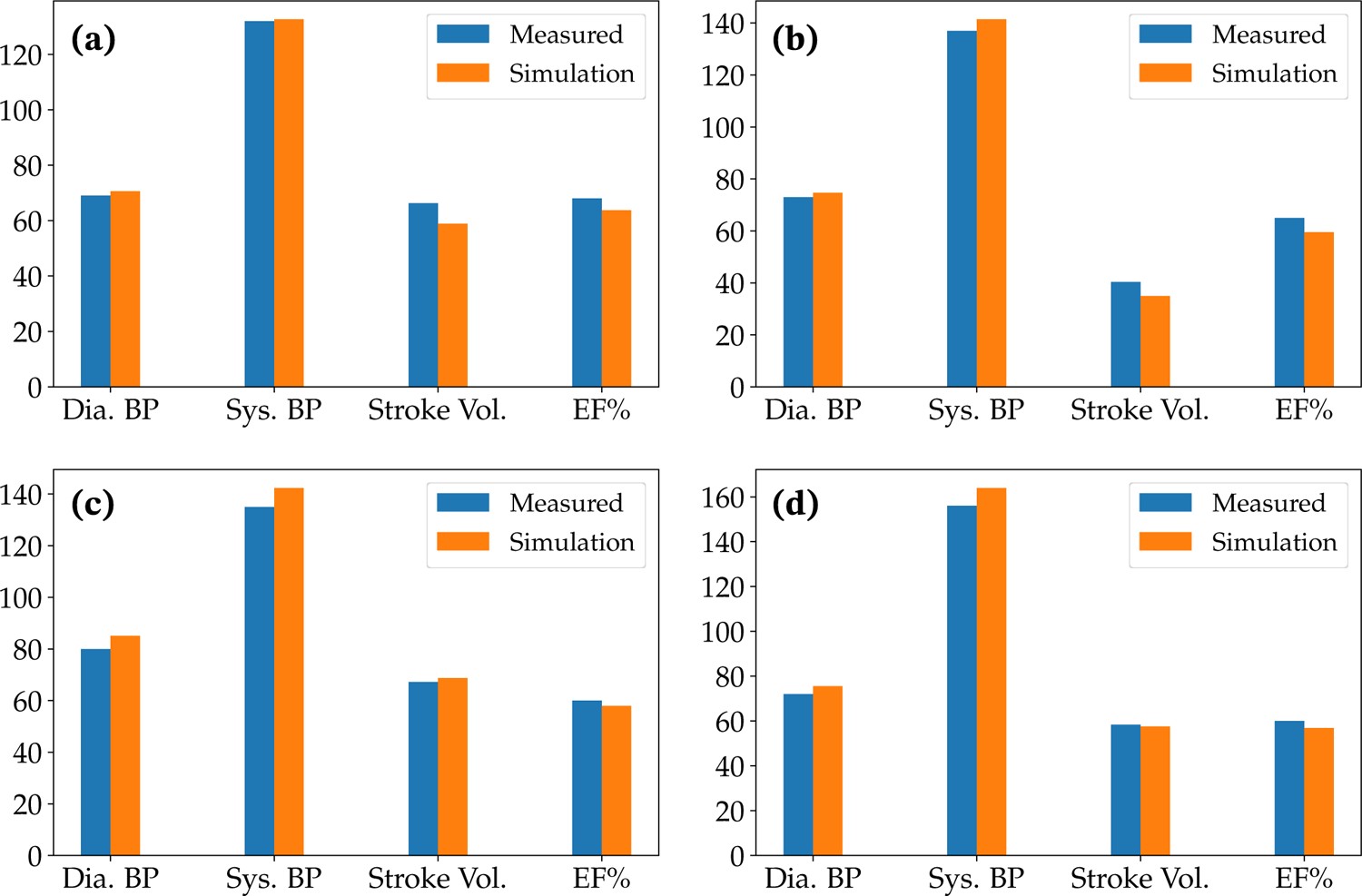
Comparison of simulated versus measured cardiac function for each patient. Figures (a), (b), (c) and (d) show data for patients 1, 2, 3 and 4, respectively. Dia. BP: Diastolic blood pressure [mmHg]; Sys. BP: Systolic blood pressure [mmHg]; Stroke vol.: Stroke volume [mL]; EF%: Ejection fraction.

Our results reveal significant differences in the flow distributions within the the coronary artery tree between models using Murray’s law versus MPI_CT_-informed boundary conditions. Figure 4 compares the simulated flow fraction at the outlet of each coronary artery with the corresponding flow fractions estimated from MPI_CT_ for each case without synthetic vascular trees. As expected for all cases informed by MPI_CT_, there was reasonable agreement between the measured flow fractions from MPI_CT_ and simulated flow fractions. The mean error in flow fractions amongst all the coronary arteries in each case utilizing boundary conditions informed by MPI_CT_ was 8.85% for case 1, 11.06% for case 2, 10.80% for case 3, and 21.04% for case 4. We note that the low-quality imaging in case 4 posed an added challenge to the parameter estimation framework due to fewer coronary arteries perfusing the LV. Some arteries were therefore associated with unfeasible/unphysiological perfusion territories and flow fractions. Moreover, this case also included lesions at the ostium of the right coronary artery and other junctions between arteries, which were not captured in the 0D surrogate used for optimization (as is the case with most common 0D and 1D surrogate models). This is discussed further in section iii.

**Figure 4:**
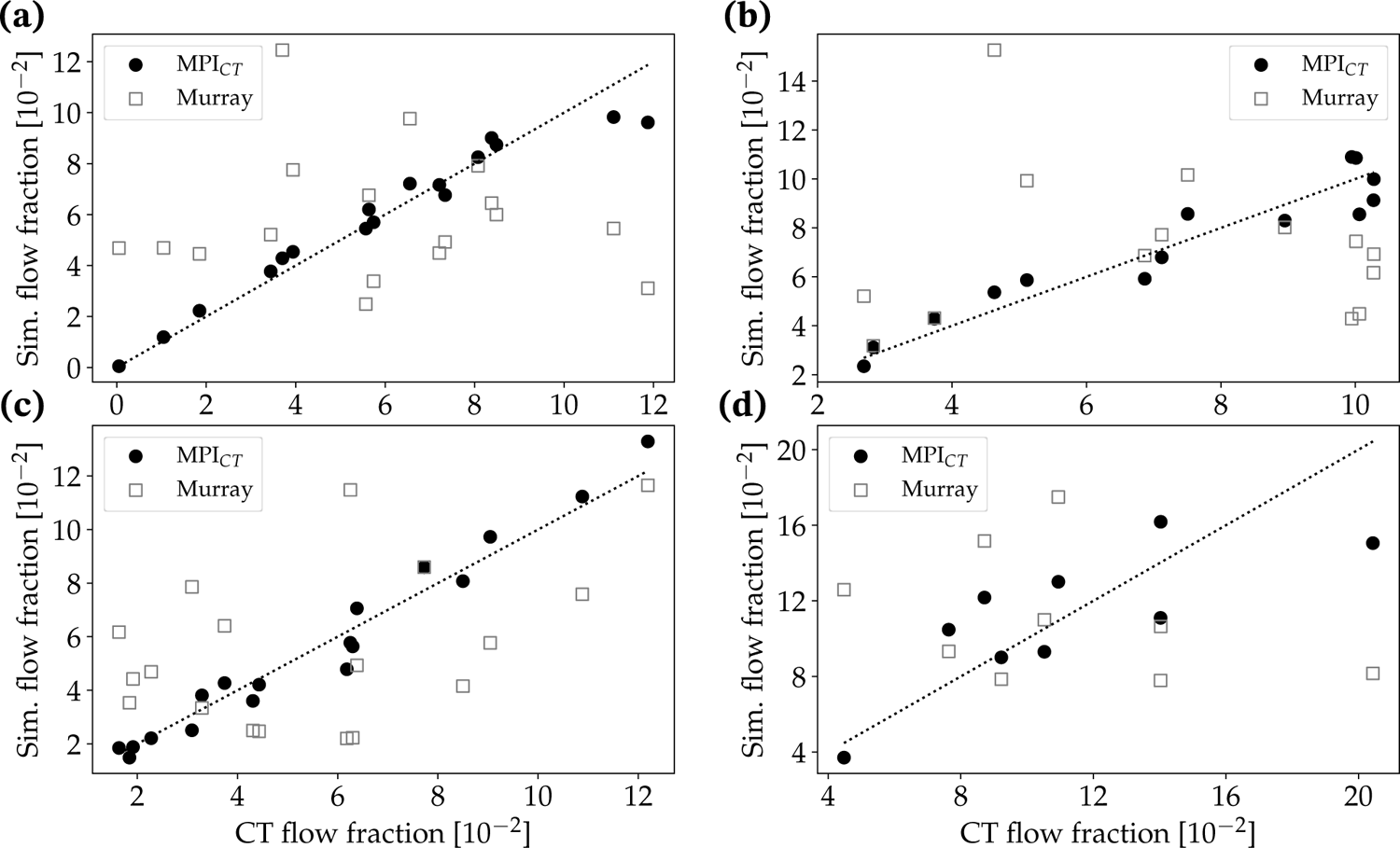
Comparison of simulated versus measured flow fraction in each coronary artery for cases without synthetic vascular trees. The simulated flow fractions are shown for cases using boundary conditions informed by MPI_CT_ as well as Murray’s law. Figures (a), (b), (c) and (d) show data for patients 1, 2, 3 and 4 respectively.

In contrast, the simulations utilizing boundary conditions based on Murray’s law showed large deviations from the measured flow distributions. The mean error between the measured and simulated flow fractions amongst all the coronary arteries was 626.72% for case 1, 50.73% for case 2, 71.55% for case 3, and 53.93% for case 4. By assessing the coronary artery branches in each case that showed the highest deviations from the measured flow distribution, we found that the branches with the largest errors were most often those affected by stenoses. However, there was no clear trend in the Murray’s law boundary conditions either over- or under-estimating the flow in branches affected by lesions.

We also compared simulated MBF in the LV (section vi) against measured MPI_CT_ on a branch-specific basis, i.e. the MBF within the regions of the LV associated with each coronary artery branch. Similar to the coronary flows, the simulated MBF distribution showed good agreement with MPI_CT_ when using the framework developed in this work, and much larger deviations when using Murray’s law-based boundary conditions (figure 5 without synthetic vascular trees). The mean territory-wise error in MBF fractions between MPI_CT_ and simulations was 3.13% for case 1, 0.78% for case 2, 7.33% for case 3 and 4.44% for case 4. In comparison, boundary conditions based on Murray’s law produced larger deviations of 17.88% for case 1, 11.32% for case 2, 26.71% for case 3 and 17.45% for case 4.

**Figure 5:**
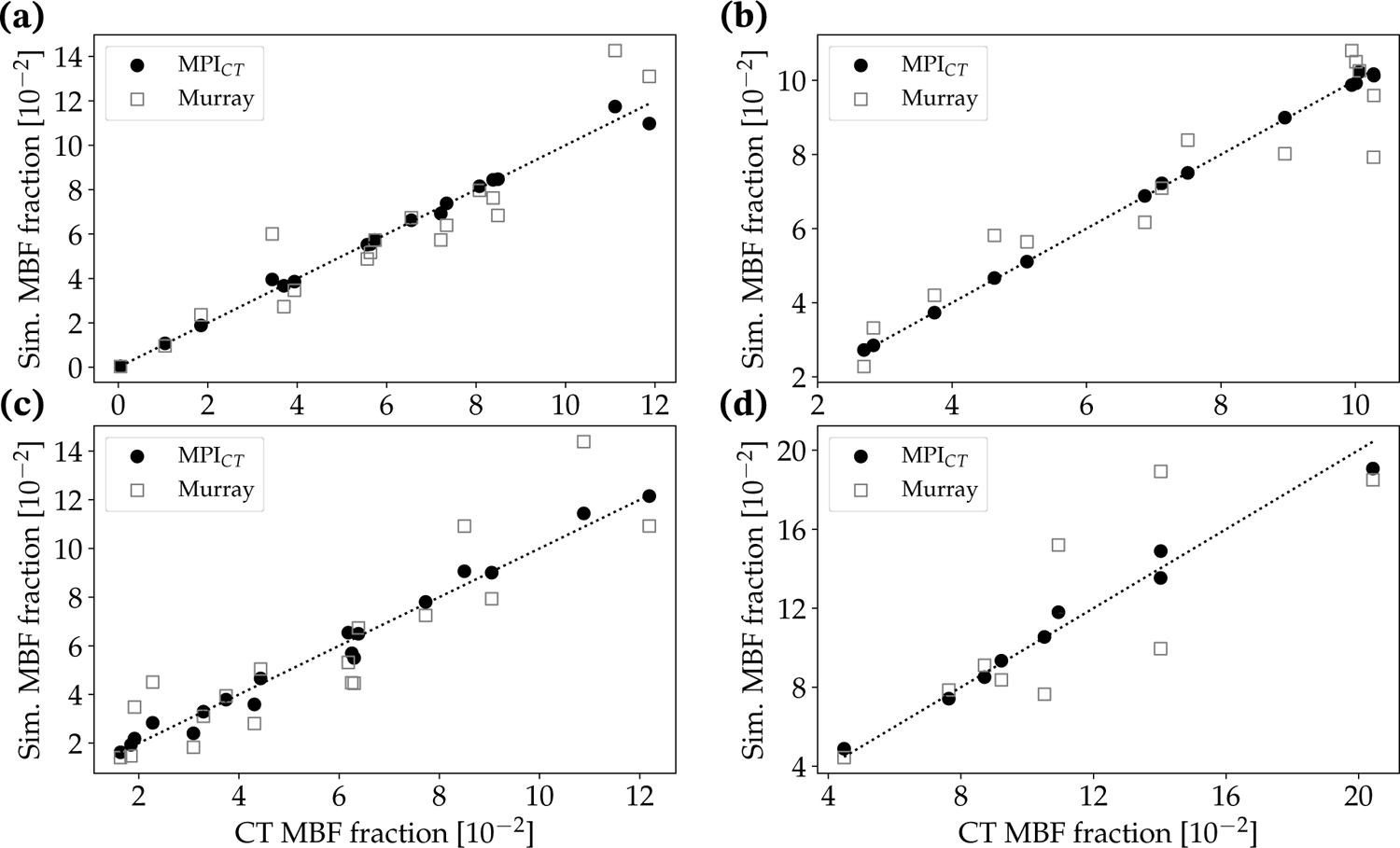
Comparison of simulated versus measured MBF fraction in each LV perfusion volume corresponding to a distinct coronary artery for cases without synthetic vascular trees. The simulated MBF fractions are shown for cases using boundary conditions informed by MPI_CT_ as well as Murray’s law. Figures (a), (b), (c) and (d) show data for patients 1, 2, 3 and 4 respectively.

Similar trends were observed for the simulations with synthetic vascular trees. Figure 6 compares the simulated coronary flow fractions at all 500 synthetic tree outlets perfusing the LV with the flow distribution measured by MPI_CT_ for all four cases. Similarly, figure 7 shows the simulated and measured MBF fractions in each vessel-specific LV perfusion territory. The mean error in flow fractions was between the simulation and MPI_CT_ was 14.48% for case 1, 12.68% for case 2, 20.32% for case 3, and 23.45% for case 4. In comparison, when using boundary conditions based on Murray’s law the mean errors in flow fractions were 130.45% for case 1, 79.77% for case 2, 79.97% for case 3 and 98.98% for case 4. The mean errors in MBF fraction when using MPI_CT_-informed simulations were 4.50% for case 1, 2.23% for case 2, 7.28% for case 3 and 6.25%for case 4. These errors increased when using Murray’s law to 21.04% for case 1, 14.30% for case 2, 32.83% for case 3 and 24.29% for case 4.

**Figure 6:**
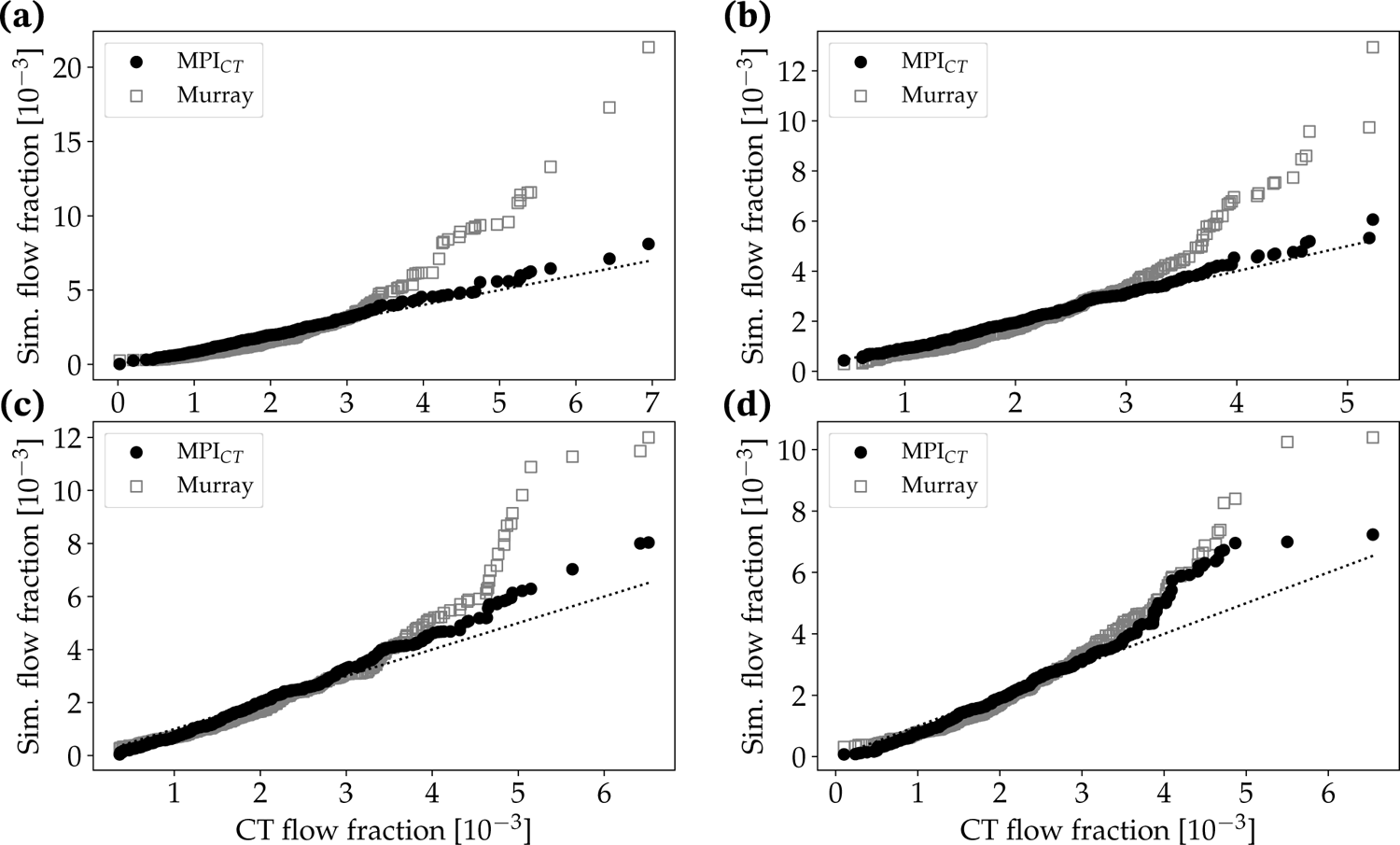
Comparison of simulated versus measured flow fraction in each coronary artery for cases with synthetic vascular trees. The simulated flow fractions are shown for cases using boundary conditions informed by MPI_CT_ as well as Murray’s law. Figures (a), (b), (c) and (d) show data for patients 1, 2, 3 and 4 respectively.

**Figure 7:**
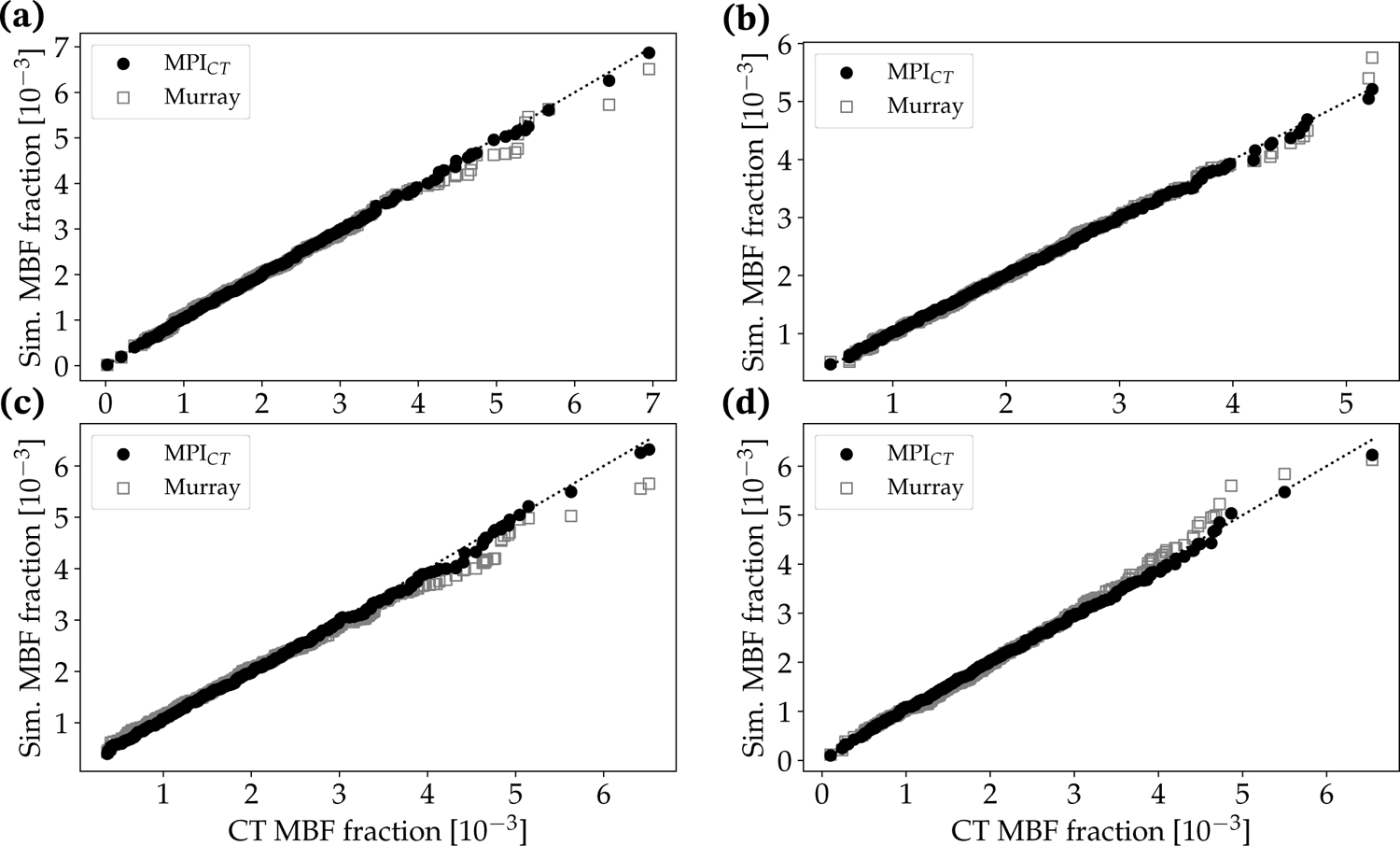
Comparison of simulated versus measured MBF fraction in each LV perfusion volume corresponding to a distinct coronary artery for cases with synthetic vascular trees. The simulated MBF fractions are shown for cases using boundary conditions informed by MPI_CT_ as well as Murray’s law. Figures (a), (b), (c) and (d) show data for patients 1, 2, 3 and 4 respectively.

The utility of augmenting computational simulations with synthetic microvascular vascular trees to model physiological MBF distributions in the LV is highlighted in figure 8. Using case 1 as an example, figure 8(a) shows the clinically imaged MBF distribution on the LV from MPI_CT_. This is compared with simulated MBF using synthetic vascular trees and Murray’s law boundary conditions in figure 8(b), MPI_CT_-informed boundary conditions in figure 8(c), and without synthetic vascular trees in figure 8(d). It is evident that the MBF distribution obtained from the model with synthetic vascular trees and MPI_CT_-informed boundary conditions best captured the clinically-imaged MBF distribution, while the Murray’s law-based simulation did not qualitatively agree with the clinical imaging. Although the simulation result without synthetic vascular trees (and with MPI_CT_-informed boundary conditions) agreed well with clinically imaged MBF on a per-vessel basis (figure 5), the small number of vessels perfusing the LV led to an unphysiological MBF distribution on the LV (figure 8d). This illustrates the utility of the hybrid image-based and synthetic vascular models used in this work, particularly in the context of simulating physiological MBF distributions.

**Figure 8:**
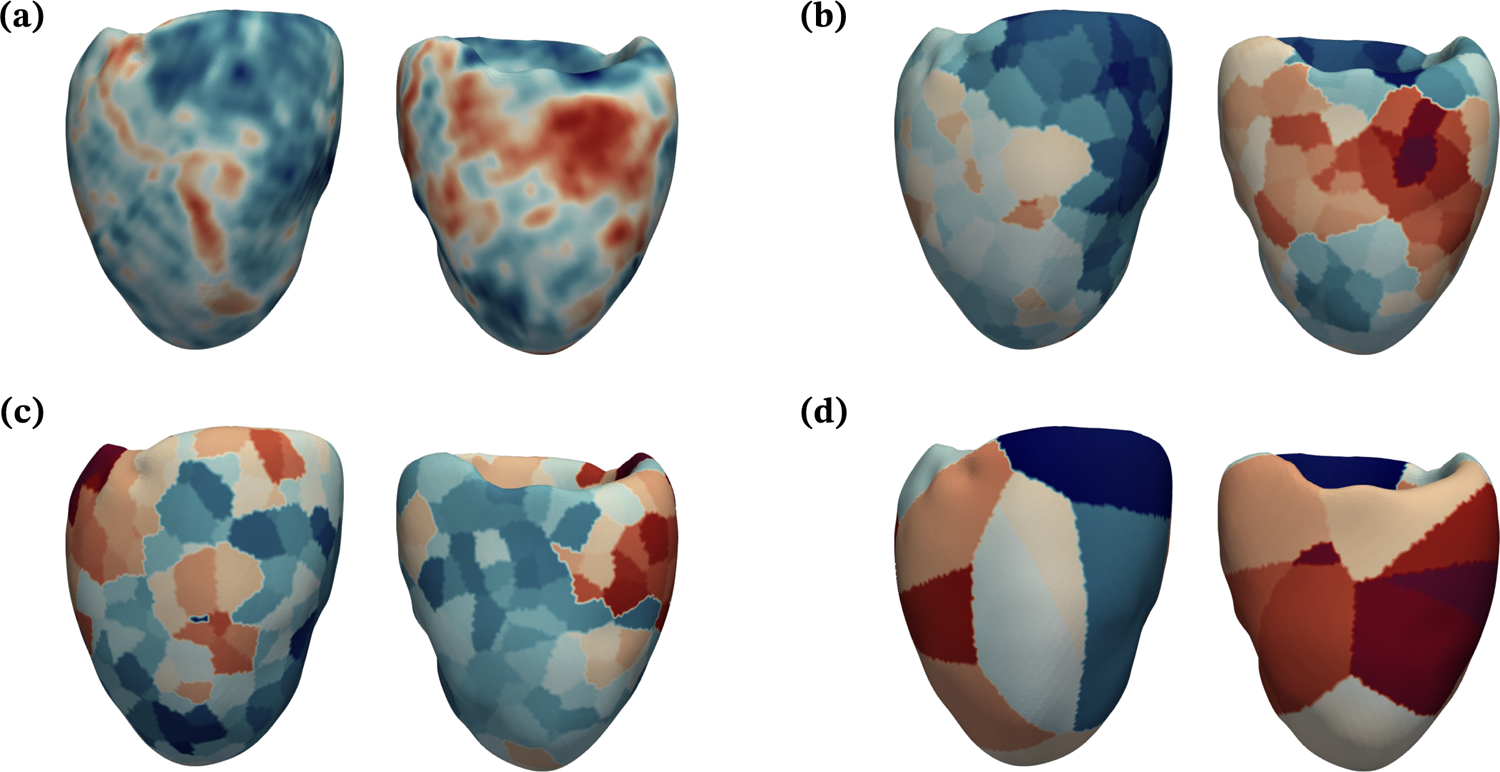
Qualitative comparison of MBF distribution on the LV for patient 1. For each sub-figure, the posterior view is shown on the left and anterior view on the right. (a) MBF from MPI_CT_. (b) MBF simulated using boundary conditions based on MPI_CT_ and synthetic vascular trees. (c) MBF simulated using boundary conditions based on Murray’s law and synthetic vascular trees. (d) MBF simulated using boundary conditions based on MPI_CT_ without synthetic vascular trees.

In current clinical practice, a widely used application of coronary hemodynamcs simulations is the non-invasive estimation of FFR using CCTA-based anatomical models [72]. We therefore compared FFR along the main stenosed branches of each case computed from our simulations using boundary conditions based on Murray’s law versus MPI_CT_ flow distributions (figure 9. We observed that the difference in FFR computed using these two methods was vessel-dependent, with the RCA exhibiting substantial differences in cases 1, 2 and 3. Differences were especially pronounced for case 1 (figure 9a), where the FFR computed using Murray’s law was below clinically-used thresholds that signify ischemic risk (FFR < 0.80 [52]) while the FFR computed using flow distributions from MPI_CT_ was above this threshold. In this case, the RCA stenosis was approximately 50% based on invasive angiography and quantitative coronary angiography (QCA), however invasive FFR measurements were not available. This is also true, although to a smaller extent, in cases 2 and 3 where the RCA had 40-60% stenoses and case 4 where the LAD had an intermediate-severity stenosis according to the angiographic readings and QCA. In comparison, both simulations based on MPI_CT_ and Murray’s law flow distributions produced FFR ≲ 0.80 for the circumflex and LAD in cases 1 and 2, the LAD in case 3, and the diagonal branch in case 4, all of which had stenoses in the range 60-70% or more, and/or diffused lesions, according to the angiographic readings and QCA.

**Figure 9:**
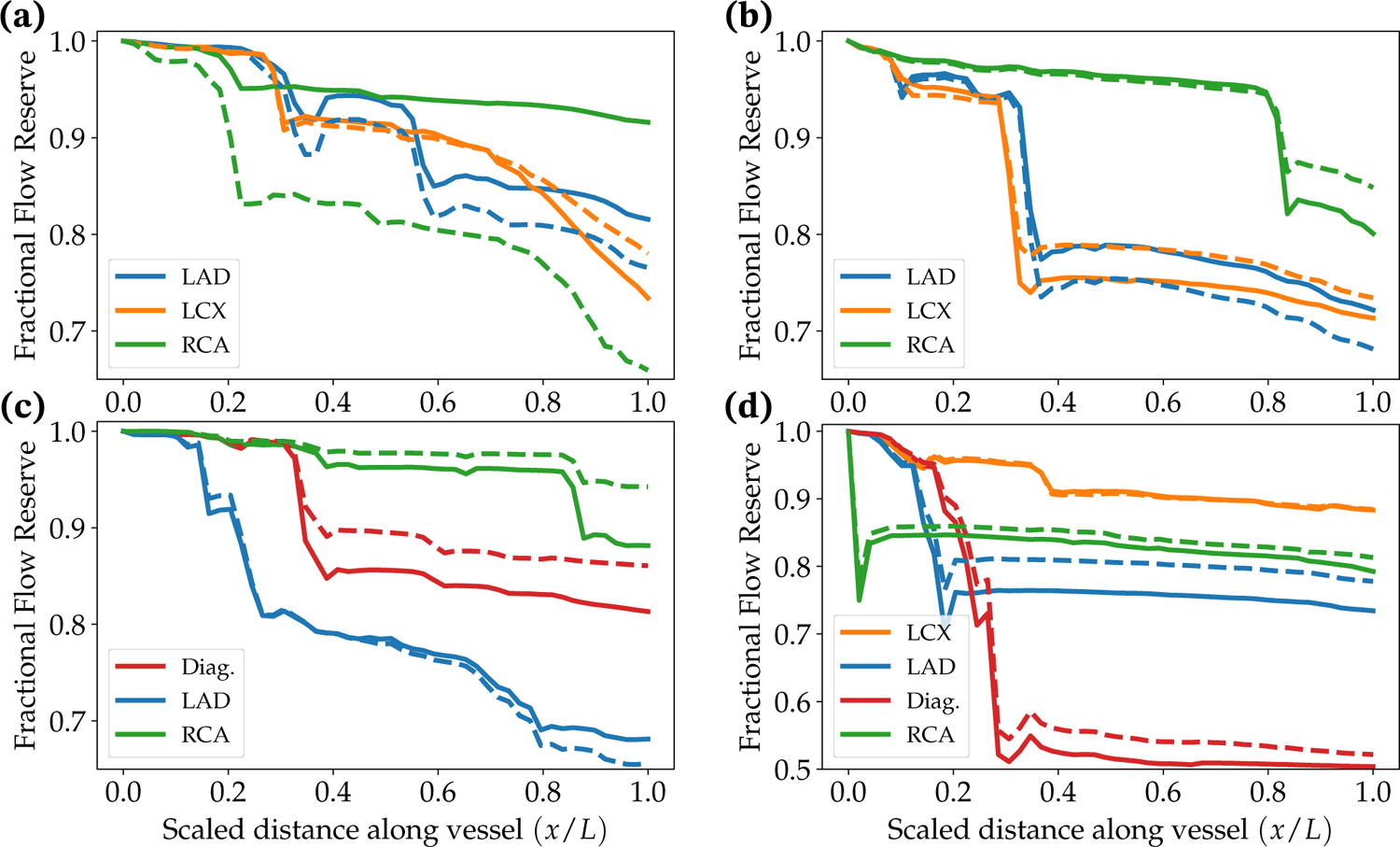
Simulated FFR using boundary conditions based on MPI_CT_ (solid lines) and Murray’s law (dashed lines). (a), (b), (c) and (d) show data for patients 1, 2, 3 and 4, respectively. For each vessel, the X-axis goes from 0 at the ostium to 1 at the outlet. LAD: Left anterior descending artery; LCX: Left circumflex artery; RCA: Right coronary artery; Diag.: Left diagonal artery.

Lastly, we compared the scaling of distal resistances at each coronary outlet obtained from the current MPI_CT_-based framework with those prescribed according to Murray’s law. This is shown for the left and right coronary artery trees for case 1 in figures 10(a) and 10(b) respectively, and for the left and right trees of case 2 in figures 10(c) and 10(d) respectively. We obtained a power-law fit between outlet resistance (*R_o_*) and coronary artery diameter (*d*) given by *R_o_ d^−^*^0.43^ (R-squared = 0.17) and *R_o_ d^−^*^0.78^ (R-squared = 0.27) for the left and right coronary artery trees for case 1, and *R_o_ d^−^*^0.23^ (R-squared = 0.10) and *R_o_ d^−^*^0.31^ (R-squared = 0.13) for the left and right trees for case 2. It is evident that there is not a good power-law fit for the coronary artery diameter-flow relationship for these cases, and the obtained scaling differs significantly from the Murray’s law relationship. This underscores the inter-patient variability in coronary flow distributions and consequently, the need to incorporate novel clinical imaging such as MPI_CT_ to personalize computational models.

**Figure 10:**
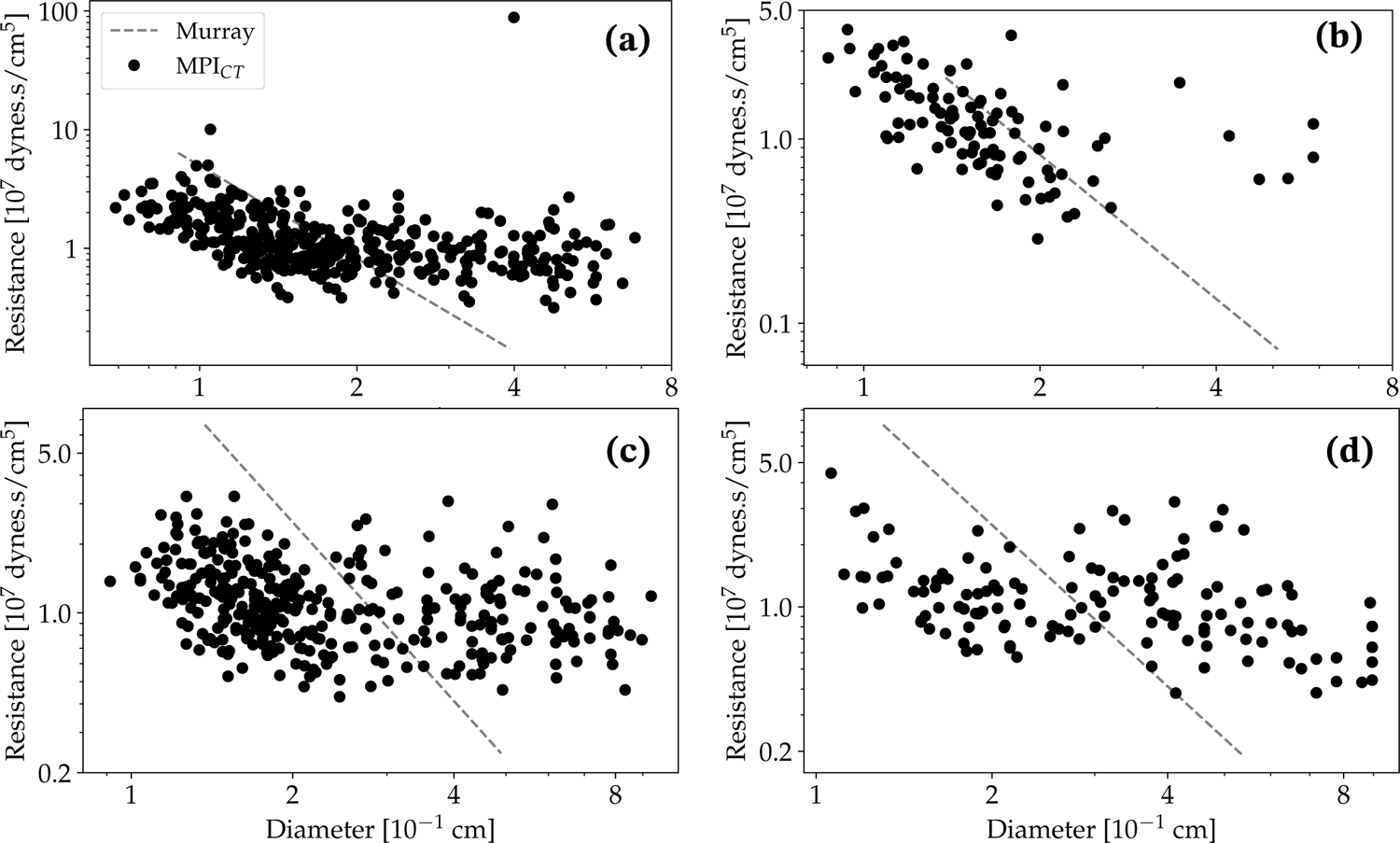
Diameter versus estimated resistance at outlets perfusing the LV based on MPI_CT_. The dashed lines show the Murray’s law scaling with exponent 2.6 for comparison. (a) and (b) show the left and right coronary tree for patient 1, and (c) and (d) show the left and right coronary tree for patient 2.

## IV. Discussion

We introduced an automated pipeline for personalizing computational models of coronary and myocardial blood flow informed by patient-specific clinical imaging and measurements. This study aimed to address two challenges in patient-specific coronary hemodynamics simulation models – the lack of patient-specific flow distributions in the coronary artery tree and inaccurate/incomplete branching patterns in image-based coronary artery anatomical models. In the following discussion, we highlight the main findings, limitations and future directions that stem from this work.

### i. Impact of MPI_CT_-informed coronary flow distributions

A major take-away from this study is that MPI_CT_-based models resulted in significantly different flow distributions than Murray’s law-based models. The largest differences in coronary artery flow between occurred distal to lesions. Moreover, our findings suggest that there is no consistent relationship across patients between the diameter of a coronary artery and its corresponding flow as measured by MPI_CT_, albeit in a small cohort. This implies that the errors in coronary flow distribution arising from Murray’s law based boundary conditions are not predictable a priori. However, this should be confirmed with larger patient cohorts and may differ in healthy patients, according to the work of van der Giessen *et al.* [78]. Crucially, the differences in flow within epicardial vessels between the models informed by MPI_CT_ and Murray’s Law affected the computation of FFR, particularly in vessels with borderline lesions of ≈ 50% stenosis. Risk stratification based on hemodynamics is particularly valuable in these lesions [73, 44], hence, the implications of these findings for the clinical use of computational models warrants more attention.

The difference in flow distributions resulting from Murray’s law and MPI_CT_ likely stems from inter-patient variability arising from coronary regulation, metabolic demand, etc., which are especially important in diseased coronary arteries. Our findings agree with previous studies that have shown significant hemodynamic differences, especially in wall shear stress, between models that used boundary conditions based on Murray’s law versus *in vivo* flow measurements [78, 62, 32]. We note, however, that these studies only included models of individual coronary arteries or single bifurcations, rather than the entire coronary artery tree simulated here.

Another reason for the inaccurate flow distributions resulting from Murray’s law-based boundary conditions is its strong dependence on the outlet diameter of arteries. In the context of image-based computational models, these diameters are derived from segmentations that are error-prone. In contrast, one can view the use of MPI_CT_-based boundary conditions as a lumped-parameter correction for inconsistencies that arise in the reconstruction of vessel lengths and lumen diameters in image-based models. For example, the resolution of clinical imaging dictates the length of each vessel visible in the image. However, this is not reflective of the actual length of the vessel, and its resultant hydrodynamic resistance or pressure drop. This therefore can be corrected for by optimizing lumped-parameter boundary condition resistances to compensate for inaccuracies in image-based segmentation. Indeed, recent work by Xue *et al.* [86], who demonstrated a framework to compute FFR from computational models informed by MPI_CT_, showed the robustness of such computational models to inaccuracies in segmenting coronary epicardial vessels.

In the context of clinical translation, a particular advantage of MPI_CT_-informed models is the ability to provide high-resolution, full coverage quantification of MBF. The ability to resolve vessel-specific MBF distributions (in contrast to lower-resolution perfusion imaging methods such as PET/SPECT) is especially valuable in the context of the framework demonstrated here. Moreover, there are clear advantages to using a single imaging modality to produce both CCTA and MPI_CT_ so that the entire framework can be integrated into a single non-invasive imaging protocol. The prognostic value of MPI_CT_ for detection of functionally significant CAD was previously demonstrated in several studies [1, 18, 33, 47], and its incorporation into computational models has substantial potential for added clinical value.

### ii. Synthetic vascular trees and multi-scale modeling

Another contribution of this work is the incorporation of the automated model personalization with multi-scale models of coronary heomdyamics. Our framework included flow in vessels smaller than the CCTA imaging resolution, microvascular perfusion in the myocardium, and LPN models of cardiac function and systemic circulation.

Our use of synthetic vascular trees appended to image-based coronary vessels is motivated by studies that have highlighted the effect of inaccurate coronary artery branching on modeled hemodynamics [82, 79]. Synthetic vascular trees provide a viable method to address this challenge even when using imaging of insufficient resolution, especially when combined with published morphometric data [26] to create realistic vascular anatomies. In addition, we demonstrated that models of myocardial perfusion that include more uniform and realistic vasculature can more accurately recapitulate clinically imaged MBF distributions. This also translates to the assimilation of data from MPI_CT_ data into computational models, i.e. higher resolution coverage of the perfusion volume by synthetic vasculature allows higher-fidelity assimilation of MPI_CT_ data into computational models.

Furthermore, our incorporation of closed-loop LPN models representing cardiac function allows us to further personalize models based on clinical measurements, rather than using literature-based boundary conditions. Such LPN models have been previously used for model-based treatment planning and to model patient-specific changes in cardiac function [64]. Finally, the coupling of the upstream hemodynamics in the coronary artery tree with the microvascular blood flow in the myocardium could enable models of coronary revascularizaton on myocardial tissue. This is infeasible in prior models of coronary hemodynamics that focus exclusively on epicardial hemodynamics [72]. In addition, this multi-scale framework opens up the possibility of studying microvascular disease using computational models.

### iii. Limitations and future work

We recognize several limitations in this study. The framework presented was deterministic, utilizing single measurements of the clinical targets for each patient to optimize the computational models. However, each clinical measurement is associated with various sources of uncertainty from the measurement protocols, patient physiology, as well as post-processing. Future work should account for uncertainties in the estimation of patient-specific parameters as well as in the model predictions; this can be performed following recent work in uncertainty quantification by our group and others [66, 67, 17]. Similarly, the sensitivity of the model to different parameters also needs to be assessed. Additionally, the dataset used here did not include clinically measured intra-coronary flow to compare with the estimated flows from MPI_CT_. While the goal is to build a non-invasive framework based entirely on MPI_CT_, comparisons with intra-coronary measurements would be valuable to increase confidence.

Another area for improvement is the complexity of the modeling utilized. While we used 3D simulations of coronary hemodynamics to compute FFR, the parameter estimation used 0D surrogate models that deviated from the true hemodynamics. In particular, 0D models do not accurately capture the flow within stenoses and at junctions between arteries. This contributed to larger errors in the flow distributions predicted from 3D models than the thresholds specified in the surrogate-based parameter estimation. Improved surrogate models will also be valuable for applying this method within time-frames that are feasible for clinical use. Another simplification was the single-compartment Darcy model we used to simulate myocardial perfusion. Previous work [22] has shown heterogeneity and anisotropy in the myocardial microvasculature, which are ignored by the assumption of constant, isotropic permeability. Moreover, the permeability was not tuned to patient-specific measurements.

This framework would also benefit from a more physiological partitioning of the myocardium into regions perfused by each coronary artery. Although there is evidence that each part of the coronary artery tree perfuses a distinct volume of the myocardium [71, 9], there is no consensus method to perform this partitioning in previous studies [62, 13, 49, 86]. However, these perfusion regions affect the simulated flow through each coronary artery. They also influence the structure of the synthetic vascular trees that extend into the myocardium from epicardial coronary arteries. While we generated vascular trees within pre-defined LV perfusion regions, future work will include methods to grow larger synthetic vascular trees that compete to perfuse physiological regions of the myocardium.

As mentioned earlier, the main goal of this work was to develop models for personalized CAD treatment planning. To that end, future work will explore applying these tools to predict the hemodynamics of patients after CABG surgery. This will involve studies in larger patient cohorts and methods to account for post-surgical vascular and cardiac adaptation.

## V. Conclusions

We introduced an automated pipeline for personalized and realistic computational models of coronary and myocardial blood flow informed by patient-specific clinical imaging and measurements. We used image-based coronary artery anatomical models augmented with synthetic vascular trees to create patient-specific anatomical models with more realistic branching. We personalized these models using flow distributions informed by MPI_CT_ and measurements of cardiac function by ultrasound. The simulation framework included three-dimensional simulations of blood flow in the coronary arteries and the left ventricle, coupled with closed-loop LPN models of distal and systemic circulation. We showed that these personalized models produced significantly different results from those based on conventionally used boundary conditions. This has important implications for the non-invasive computation of clinically-relevant metrics, such as FFR and MBF, from CFD simulations.

## Data Availability

All computational models built for this study have been anonymized and made publicly available through the Vascular Model Repository (www.vascularmodel.com).

## Acknowledgments

This work was supported by NIH grant R01HL141712 and the Additional Ventures Cures Collaborative. Z.A.S. was funded by the NSF Graduate Research Fellowship Program (DGE-1656518). High performance computing resources were provided by XSEDE and the Stanford Research Computing Center.

## VII. Code availability

The patient-specific modeling pipeline and computational fluid dynamics solvers are part of the SimVascular open-source project (www.simvascular.github.io). Image analysis and postprocessing was also performed using the open-source tools specified in section ii.

